# The potential effect of the African population age structure on COVID-19 mortality

**DOI:** 10.1101/2020.05.19.20106914

**Authors:** Fabrice Mougeni, Ance Mangaboula, Bertrand Lell

## Abstract

Currently (mid May 2020), most active cases of COVID-19 are found in Europe and North America while it is still in the initial phases in Africa. As COVID-19 mortality occurs mainly in elderly and as Africa has a comparably young population, the death rates should be lower than on other continents.

We calculated standardised mortality ratios (SMR) using age-specific case fatality rates for COVID-19 and the age structure of the population of Africa and of other continents. Compared to a European or Northern American population, the standardised mortality ratio was only 0.22 and 0.25, respectively, corresponding to reduction of deaths rates to a quarter. Compared to the Asian and Latin American & Caribbean population, the SMR was 0.43 and 0.44, respectively, corresponding to half the death rate for Africa.

It is useful to quantify the isolated effect of the African age-structure on potential COVID-19 mortality for illustrative and communication purposes, keeping in mind the importance of public health measures that have been shown to be effective in reducing cases and deaths. The different aspect of age pyramids of a European and an African population are striking and the potential implications for the pandemic are often discussed but rarely quantified.

## Introduction

Currently (mid May 2020), most active cases of COVID-19 are found in Europe and North America while it is still in the initial phases in Africa. It is unclear what death rates can be expected for this continent. On one hand, African health care systems are weak and therefore many severely ill patients will not be able to receive ventilation and other high-end health care in intensive care units. On the other hand, COVID-19 mortality occurs mainly in elderly and as Africa has a comparably young population, the proportion of deaths among the population should be lower than on other continents.

As age-specific case fatality rates for COVID-19 are available, and the age structure of the population of Africa and of other continents is known, it is possible to calculate the comparative effect of age using methods known from indirect age adjustment.

## Methods

Age-specific case fatality rates (CFR) published in March 2020 were used.(1) These were based on data mainly from China and were calculated using a model correcting for biases such as the preferential detection of severe cases early in the epidemic, and the delay in time from detection to outcome. Although there since have been over ten times the number of cases since these calculations were published, they are still likely to be best currently available estimates and no updates on age-specific case fatality rates are available. Two sets of CFR were used and compared, the crude CFR and the CFR adjusted for censoring, demography and under-ascertainment. The 2020 population estimates of the “United National World Population Prospects” were used to find the proportion of each population within each age group.(2)

Indirect age adjustment was performed by multiplying the case fatality rate with the proportion of the population within an age group. The ratio of the sums, which corresponds to the standardised mortality ratio (SMR) in indirect age adjustment show the relative number of deaths in equally sized populations. We use the term standardised mortality ratio, as it closely corresponds to indirect age adjustment procedure. However, we use it to calculate relative number of deaths rather than comparing predicted versus actual deaths. The formula used for the calculation of the SMR is as follows:

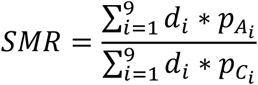

The proportion of population within each of the 9 age groups are designated *p_Ai_* for Africa and *p_Ci_* for comparator populations. The age-specific case-fatality rate is designated *d_i_*. Confidence interval were calculated with a bootstrap method in R using the ‘rsample’ library. (3)

## Results

The crude and adjusted CFR and the proportion of population in each age group for 5 different regions are shown in table 1.

**Table 1:** CFRs and proportion of African and other populations within each age group

| Age group<br>(yrs) | CFR (%)<br>crude | CFR (%)<br>adjusted | Africa | Europe | Northern<br>America | Asia | Latin Am.<br>Caribbean |
| --- | --- | --- | --- | --- | --- | --- | --- |
| 0–9 | 0 | 0.0026 | 0.285 | 0.107 | 0.118 | 0.157 | 0.159 |
| 10–19 | 0.182 | 0.0148 | 0.222 | 0.105 | 0.126 | 0.155 | 0.162 |
| 20–29 | 0.193 | 0.0600 | 0.168 | 0.112 | 0.139 | 0.155 | 0.164 |
| 30–39 | 0.237 | 0.146 | 0.127 | 0.141 | 0.135 | 0.155 | 0.152 |
| 40–49 | 0.443 | 0.295 | 0.087 | 0.139 | 0.123 | 0.133 | 0.129 |
| 50–59 | 1.30 | 1.25 | 0.057 | 0.139 | 0.128 | 0.115 | 0.105 |
| 60–69 | 3.60 | 3.99 | 0.035 | 0.123 | 0.117 | 0.077 | 0.072 |
| 70–79 | 7.96 | 8.61 | 0.016 | 0.081 | 0.073 | 0.038 | 0.039 |
| ≥80 | 14.8 | 13.4 | 0.004 | 0.053 | 0.040 | 0.015 | 0.019 |

The standardized mortality ratios comparing the African and other populations are shown in table 2.

**Table 2:** Standardised mortality ratios between African and other populations

| Populations | SMR (95% CI)<br>crude CFR | SMR (95% CI)<br>adjusted CFR |
| --- | --- | --- |
| Africa vs. Europe | 0.24 (0.20; 0.25) | 0.22 (0.18; 0.22) |
| Africa vs. Northern America | 0.28 (0.24; 0.28) | 0.25 (0.24; 0.25) |
| Africa vs. Latin Am. & Caribbean | 0.47 (0.46; 0.47) | 0.43 (0.43; 0.43) |
| Africa vs. Asia | 0.48 (0.48; 0.48) | 0.44 (0.44; 0.44) |

## Discussion

COVID-19 mortality is strongly age-dependent and Africa has a young population compared to other regions. We show that its isolated effect is quite strong in comparison to Europe or Northern America, and one can expect only around a fourth of the death rate simply due to the age effect. In comparison, Latin American and Asian populations have a higher proportion in younger age groups, but the effect is still clear when compared to Africa, with a reduction of around 50%.

The number of COVID-19 deaths varies considerably from country to country and from district to district within countries, due to differences in e.g. public health measures, and restrictions taken by the authorities. Our results should therefore not be interpreted as predictive.

Authors of mathematical models predicting COVID-19 mortality in Africa acknowledge the strong effect of age in the African population.(4,5) However, it is useful to quantify the isolated effect of the African age-structure on potential COVID-19 mortality for illustrative and communication purposes. The different aspect of age pyramids of a European and an African population are striking and the potential implications for the pandemic are often discussed but rarely quantified.

## Data Availability

All data used are publicly available from the sources cited in the text

## References

1. Verity R, Okell LC, Dorigatti I, Winskill P, Whittaker C, Imai N, et al. Estimates of the severity of coronavirus disease 2019: a model-based analysis. Lancet Infect Dis. 2020 Mar 30;

2. World Population Prospects - Population Division - United Nations [Internet]. [cited 2020 May 19]. Available from: https://population.un.org/wpp/Download/Standard/Population/

3. Choquet D, L’Ecuyer P, Léger C. Bootstrap confidence intervals for ratios of expectations. ACM Trans Model Comput Simul. 1999 Oct;9(4):326–48.

4. Diop BZ, Ngom M, Pougue Biyong C, Pougue Biyong JN. The relatively young and rural population may limit the spread and severity of Covid-19 in Africa: a modelling study. medRxiv. 2020 May 3;2020.05.03.20089532.

5. van Zandvoort K, Jarvis CI, Pearson C, Davies NG, Russell TW, Kucharski AJ, et al. Response strategies for COVID-19 epidemics in African settings: a mathematical modelling study. medRxiv. 2020 Jan 1;2020.04.27.20081711.

